# Combining explainable machine learning, demographic and multi-omic data to identify precision medicine strategies for inflammatory bowel disease

**DOI:** 10.1101/2021.03.03.21252821

**Authors:** Laura-Jayne Gardiner, Anna Paola Carrieri, Karen Bingham, Graeme Macluskie, David Bunton, Marian McNeil, Edward O. Pyzer-Knapp

## Abstract

Inflammatory bowel diseases (IBDs), including ulcerative colitis and Crohn’s disease, affect several million individuals worldwide. These diseases are heterogeneous at the clinical, immunological and genetic levels and result from a complex interaction between the host and environmental factors. Investigating drug efficacy in cultured human fresh IBD tissues can improve our understanding of the reasons why certain medications are more or less effective for different patients.

We propose an explainable machine learning (ML) approach that combines bioinformatics and domain insight, to informatively integrate multi-modal data to predict inter-patient specific variation in drug response. Using explanation of our models, we interpret the models’ predictions inferring unique combinations of important features associated with human tissue pharmacological responses. The inferred multi-modal features originate from multi-omic data (genomic and transcriptomic), demographic, medicinal and pharmacological data and all are associated with drug efficacy generated by a preclinical human fresh IBD tissue assay.

To pharmacologically assess patient variation in response to IBD treatment, we used the reduction in the release of the inflammatory cytokine TNFα from the fresh IBD tissues in the presence or absence of test drugs, as a measure of drug efficacy. The TNF pathway is a common target in current therapies for IBD; we initially explored the effects of a mitogen-activated protein kinase (MAPK) inhibitor on the production of TNFα; however, we later show the approach can be applied to other targets, test drugs or mechanisms of interest. Our best model was able to predict TNFα levels from a combination of integrated demographic, medicinal and genomic features with an error as low as 4.98% on unseen patients. We incorporated transcriptomic data to validate and expand insights from genomic features. Our results showed variations in drug effectiveness between patients that differed in gender, age or condition and linked new genetic polymorphisms in our cohort of IBD patients to variation in response to the anti-inflammatory treatment BIRB796 (Doramapimod).

Our approach models drug response in a relevant human tissue model of IBD while also identifying its most predictive features as part of a transparent ML-based precision medicine strategy.

## Introduction

Precision medicine has become a widely recognised and desirable medical model for its ability to stratify patients into different groups based on their susceptibility to a particular disease or their response to a specific drug [1]. The personalisation of medical decisions and the recommendation of interventions or treatments that are tailored to the individual allows patients to receive appropriate treatment more rapidly, which improves their quality of life and can reduce rising demands for health care support. Precision medicine has the potential to transform the prediction of disease progression, and therefore aid its possible prevention, and to inform precise and targeted therapies [2].

If we consider drug development specifically, a major challenge is how to effectively implement precision medicine strategies earlier in the drug development process. Stratifying patients into subpopulations at a late stage i.e., during or after demographic trials, comes with an associated high-cost burden, with approximately 70% of the cost of drug development attributed to the clinical stage [3]. If patient stratification is found to be necessary to achieve the required efficacy, this high cost/late-stage approach could have a major impact on the commercial viability of a drug due to the smaller than expected target patient population [4]. Early understanding of the target patient population at the pre-clinical stage would help address this problem by allowing the creation of earlier/more accurate economic models and would also allow a more targeted and streamlined approach to be taken during the demographic phase of development. Moreover, a failure to stratify patients prior to clinical trials likely contributes to the high failure rate during Phase II/III trials, where a key problem is a failure to reach endpoints for efficacy using an “all-comers” clinical trial.

When selecting preclinical models for use in precision medicine strategies, the most important feature of the selected model(s) is that the efficacy readout is translatable to the demographic outcome. In-line with this requirement, confirming the pharmacological activity of a drug in the target tissue has been shown to be an excellent predictor of successful demographic trials [5-7]. Human fresh tissues that reflect the native biology of disease are therefore increasingly being used during preclinical drug development to meet this aim of improving the prediction of efficacy in clinical trials. As such, here, we developed an explainable AI workflow that combines multi-omic data, demographic data, medicinal data and pharmacology data, all derived from a preclinical fresh human tissue assay, to predict patient-specific drug responses and to facilitate clinical trial precision medicine strategies (Figure 1).

**Figure 1.**
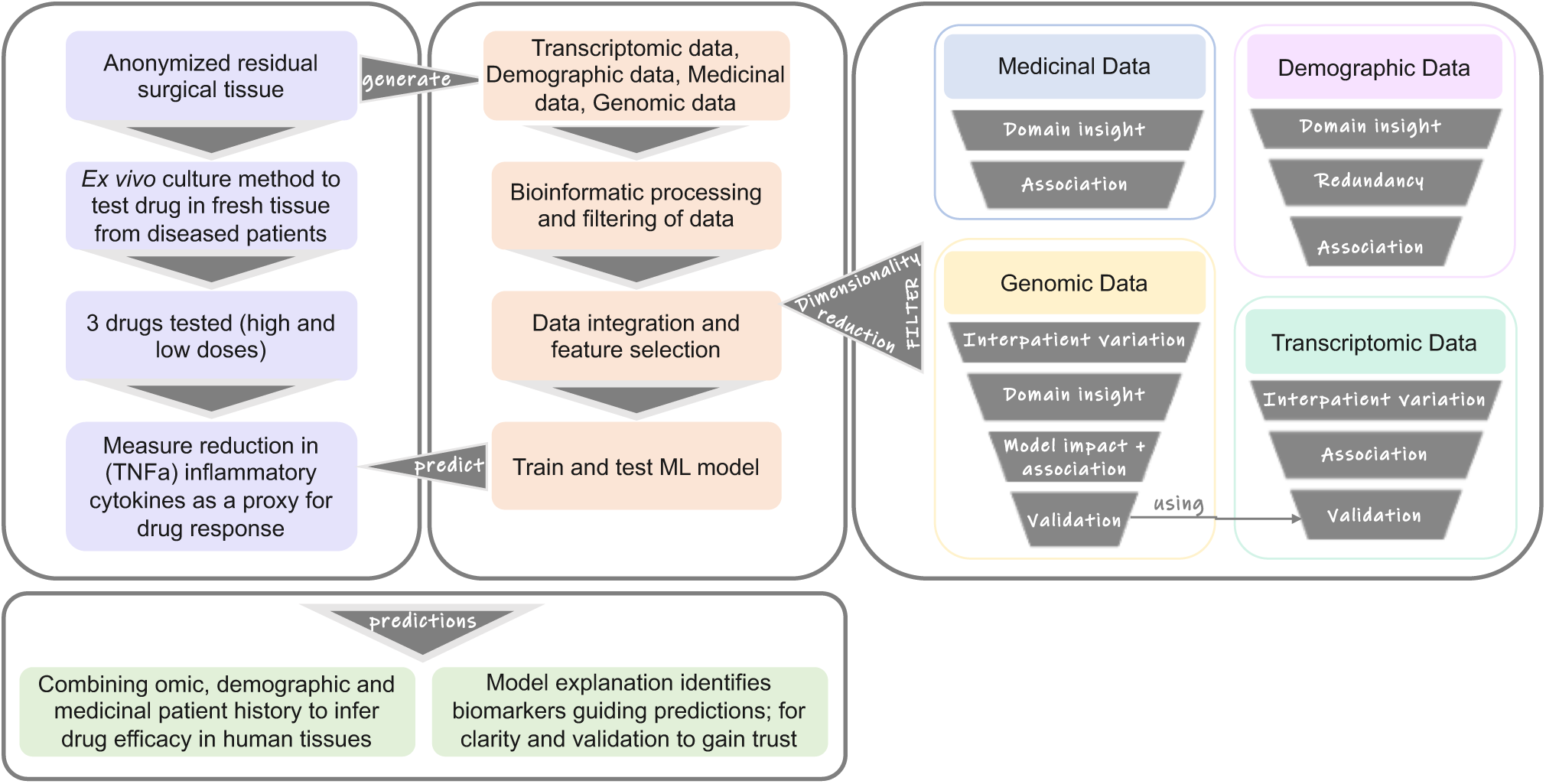
Schematic representation of the study. Detailing (to the left) the steps undertaken to generate datasets, process these datasets, build and train ML models to make predictions and the interpretation of those predictions. Detailing (to the right) the different approaches, in order of usage where possible, which we used for dimensionality reduction of the medicinal, demographic, genomic and transcriptomic feature sets that were used to train our models and provide explanations for the predictions. Ultimately models were trained to predict the TNFα level or inflammatory response after compound treatment on tissues.

We focus on drug development for inflammatory bowel diseases (IBDs) for the two primary conditions: ulcerative colitis (UC) and Crohn’s disease. UC and Crohn’s disease are long-term conditions that involve inflammation of the gut. UC only affects the colon (large intestine), while Crohn’s disease can affect any part of the digestive system, from the mouth to the anus. In 2017, there were 6·8 million cases of IBD globally [8]. The most widely held hypothesis on the pathogenesis of IBD is that overly aggressive acquired (T cell) immune responses to a subset of commensal enteric bacteria develop in genetically susceptible hosts, and environmental factors precipitate the onset or reactivation of disease [9]. Currently there is no cure for either disease and people with IBD will typically need treatment throughout their lives. As such, we need to understand why certain medications are more or less effective for different patients and relate this to their demographic information and genetic makeup [10]. To investigate the level of patient stratification that can be achieved from a preclinical study, the commonly used ‘all comers’ clinical trial patient recruitment policy was mimicked with no donor inclusion criteria other than a clinical diagnosis of UC or Crohn’s [11]. To pharmacologically assess patient variation in responsiveness to treatment, the reduction in the inflammatory cytokine TNFα in the presence or absence of test drugs was used as the measure of drug efficacy.

TNFα is powerful proinflammatory cytokine that is a key mediator in inflammatory diseases [12]. The main source of TNFα is from activated cells from the monocyte lineage e.g macrophages, however release has been shown in other cell types. In IBD in particular, TNFα has been shown to be a key signalling molecule and disease driver [13-15]. This has led to TNFα becoming a key target for IBD drug treatments, most notably with the development and introduction of therapeutic anti-TNF antibody treatments in the last 10-15 years.

Included in this study were 3 different test drugs. Two of these (5-ASA and prednisolone) are standard of care treatments for IBD which are routinely prescribed as first line treatments when patients first present with IBD [16-17]. The third drug (doramapimod, also known as BIRB796) is a drug that is not used clinically for IBD; however, it was developed for use in IBD and underwent clinical trials for use in Crohn’s disease. The rationale for including BIRB796 in the study was two-fold: firstly, as 5-ASA and prednisolone are widely used in IBD patients, it is likely that the majority of the patient samples used in our organoculture assay would already have been exposed to these test articles. Since this may have impacted their responsiveness *in vitro*, we decided to also include a drug with a novel mechanism that patients would not have been exposed to. Secondly, although BIRB796 was developed for the treatment of inflammatory diseases including IBD, it failed to progress beyond Phase 2 clinical trials due a lack of efficacy. It is therefore an example of a drug which may have benefitted from a preclinical patient stratification strategy, such as we are describing. The mechanisms of action of 5-ASA, prednisolone and BIRB796 are all thought to interact with TNFα via peroxisome proliferator-activated receptor gamma (PPAR-γ) [18], nuclear factor kappa light chain enhancer of activated B cells (NF-κB) [19] and MAPK [20], respectively. TNFα was therefore deemed to be both a disease and drug mechanism relevant marker of efficacy to utilise in this study. As all human tissue organoculture data was normalised against a patient-matched non-treated control group, the degree by which TNFα levels were reduced was interpreted as a better response to the test drug.

The individual patient organoculture assay responses were then matched with multi-omic, demographic and medicinal data of the same patients/tissues to predict patient-specific drug response, and to identify the integrated feature profiles of the patients most likely to respond to the treatment. In light of the recent advances in omic technologies and the falling cost of sequencing, it has become a more realistic aim to enable precision medicine using the routine analysis of a patient’s genetic information or genome and to combine this with other demographic information to personalise medical interventions. Furthermore, it is hypothesised that the integration of multiple sources of omic data (multi-omic) will create a more holistic picture of a disease under investigation, particularly when used alongside demographic data or other data types e.g., medical images [21]. Such heterogenous or multi-modal data integration remains one of the major challenges facing precision medicine today, where there is no widely adopted best practice methodology [22], and where biological knowledge is needed (but typically lacking) to guide integrative methods. We propose that Artificial Intelligence (AI), guided by domain knowledge, has the potential to facilitate heterogenous data integration and to offer actionable insights into medical aspects from disease progression to drug development [23].

To this end, we combine machine learning (ML), bioinformatics and domain insight, to allow the processing and informative integration of features derived from diverse multi-modal data types. These data include multi-omic data (genomic and transcriptomic), demographic data, medicinal data (prescribed medicines) and pharmacology data (functional experimental assays), all derived from a preclinical fresh human tissue assay. Features derived from this data, that are inputted into predictive models include, for example, SNPs from genomic data or patient age from demographic data. Our workflow comprised bioinformatics, feature selection, machine learning (ML) models and an explainable AI algorithm. Explainable AI helps us to understand the predictions made by ML models and offer insights into the predicted phenotype. Only recently has explainable AI been applied to diverse single omic datasets [24-25], but the potential of this to enable precision medicine has yet to be fully exploited. Here our aim, for IBD patients, is to predict patient-specific drug response and derive insights into the potential biological (genetic or otherwise) basis of this variation in response. Importantly, we use explainable AI to identify unique combinations (or profiles) of important features that are associated with drug response in human fresh tissue assays. Important features consist of those optimal combinations that led to the most accurate results when predicting drug response. These profiles could be used to analyse inter-individual drug response and to stratify patients for personalised drug treatment or to inform targeted clinical trials during drug discovery.

## Results and Discussion

### Prediction of TNFα level (drug response) from demographic information

For the processed demographic information for the 25 patients, we initially excluded features using domain insight; alcohol history (since this was missing for a large proportion of donors or labelled too ambiguously for confident interpretation), and supplier region (since this is largely uninformative with regard to the phenotype we are predicting) (see Methods, Figure 1). We next removed the redundant feature ethnicity since all the donors were Caucasian or else unknown. The remainder of the demographic features were investigated using Spearman’s correlation (Figure 2). All features were found to be correlated with response (measured TNFα level) to one or more of the compounds at a correlation |rs| > 0.3 except for smoking history. This could be a result of the inconsistent and incomplete information provided for smoking history, as such this feature was removed. Finally, we noted that resection area was highly correlated with condition, making resection area a largely redundant feature, so it was also removed from the analysis.

**Figure 2.**
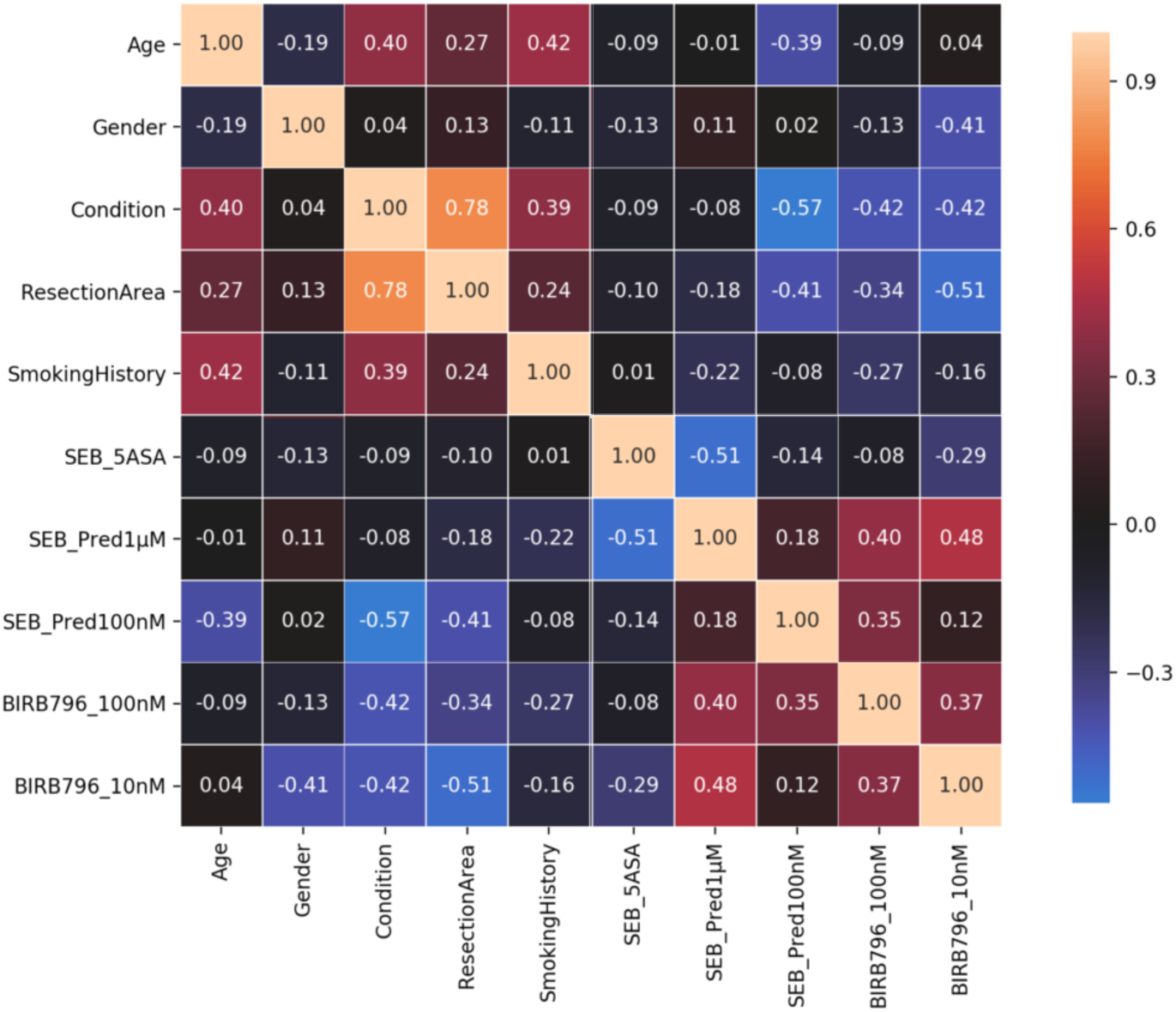
Spearman Correlation of demographic information with TNFα response per drug. To compute the Spearman correlation for binary features we used the following encoding; condition Crohn’s/ulcerative colitis = 0/1, gender female/male = 0/1, resection area colon/ileum = 0/1. See section demographic feature preparation for ML in Methods for more details.

To quantify the impact of feature selection, in particular the impact of our usage of correlation to remove additional features, we selected one of our five drug/dose combinations as an example (BIRB796 at 10nM) and compared the predictive capability of a range of ML models trained with and without the demographic features smoking history and/or resection area (see Methods). The removal of smoking history and/or resection area either reduced the predictive error or did not have a significant impact on the predictive error overall, which further justified their removal (Figure S1). Here we measured predictive error on our test dataset that represents patients that were not seen by the model during training and used solely to gauge its accuracy on new datapoints. Predictive error was quantified using the mean absolute error (MAE) that represents the average of the absolute differences between the predictions and the actual observations.

### Incorporating medicinal information for the prediction of drug response

To process the medicinal information that was available for the donors we firstly used domain insight to collapse multiple representations of the same drug due to different naming conventions, this collapsed 61 medicines down to 53. We labelled patients individually as not receiving medicine (0) or receiving medicine (1) for each of the 53 medicines and performed Spearman’s correlation, enabling the selection of 12 medicines overall that correlated with response to one or more of the tested compounds/doses at a level |rs| > 0.3 (Table 1).

**Table 1.** Medicinal features correlated with drug response at |rs| >=0.3 (Spearman’s)

| 5-ASA | Pred1uM | Pred100nM | BIRB796_100nM | BIRB796_10nM |
| --- | --- | --- | --- | --- |
| Nomeds | Adalimumab | Azathioprine | Rivaroxaban | Ranitidine |
|  |  | Mezarant | Ranitidine | Rivaroxaban |
|  |  |  | Lyrica | Lyrica |
|  |  |  | Amitriptyline | Levothyroxine |
|  |  |  | Azathioprine | Amitriptyline |
|  |  |  | Tapentadol | Prednisolone |
|  |  |  |  | Asacol |
|  |  |  |  | Tapentadol |

To quantify the benefit of adding the entire set of medicines as features to our existing demographic feature set – scenario a, we compared this to the impact of selecting only correlated medicines – scenario b. We used BIRB796 at 10nM to highlight our comparison of the predictive capability of the derivative models. Figure 3 shows that the addition of medicinal information, in both scenarios (Figure 3a or 3b), decreased the predictive error of the models compared to those obtained using only the demographic information (see Figure S1). Moreover, Figure 3 shows that including only the correlated medicines to BIRB796 provided a predictive advantage compared to including all the 53 medicines in our feature set. As such, our best “demographic+medicinal” feature set incorporated the demographic features age, gender and condition and the correlated medicinal features for BIRB796 shown in Table 1. The best performing ML model with our best feature set was KNN with a median MAE of ∼4.49% over 10-fold cross validation (Figure 3b).

**Figure 3.**
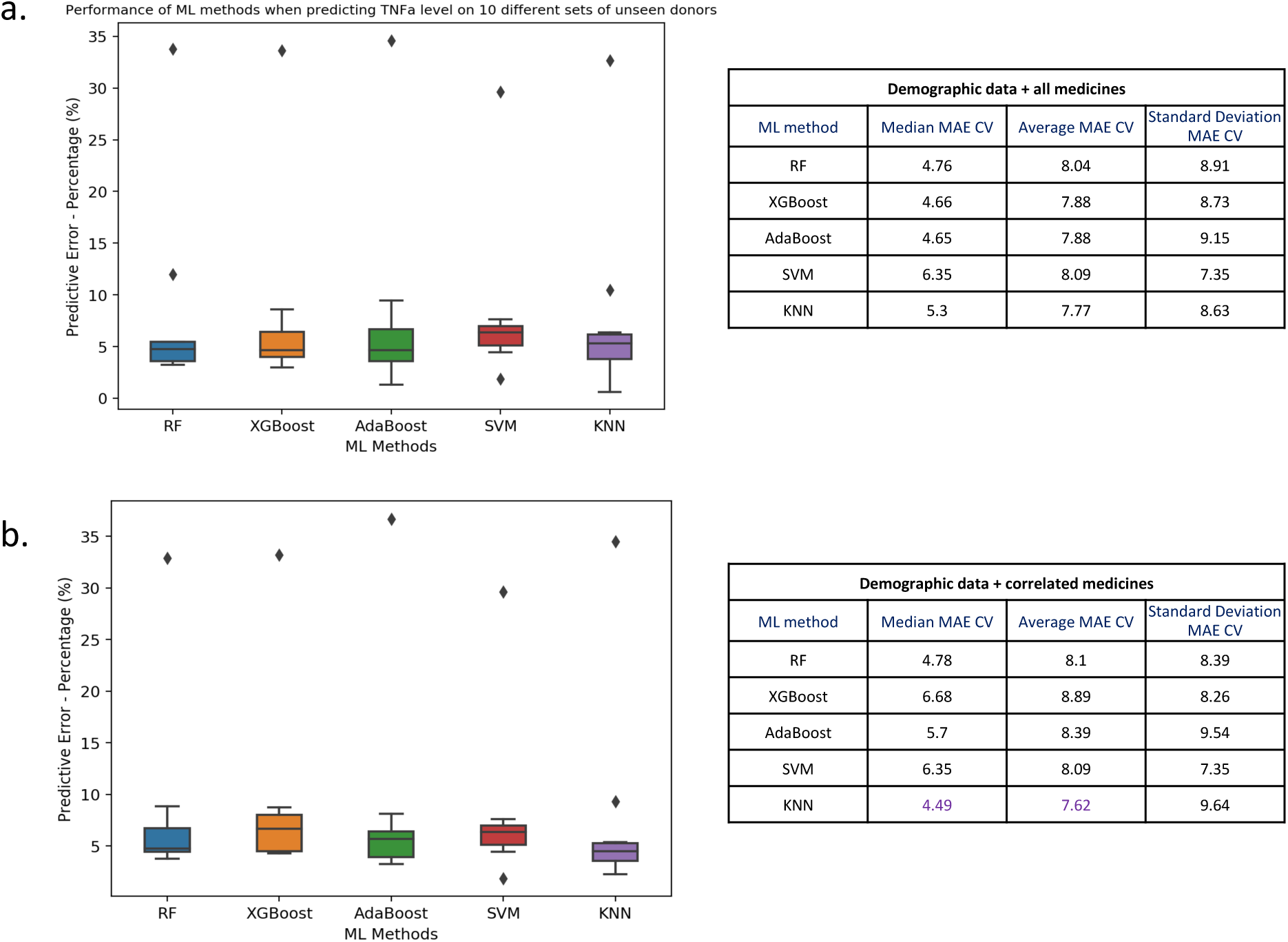
Comparison of ML model error rates for the prediction of BIRB796 (10nM) drug response for different combinations of demographic and medicinal features. Here we show box plots (left) of mean absolute error (MAE) values (as percentages) computed during 10-fold cross validation. The horizontal line in each boxplot is the median of the MAE over 10 folds, where each of the test folds has 3 randomly chosen patients. Note all target drug responses have been normalized on a scale of 0-1 and here we show percentages of MAE values. On the right we report median, average and standard deviation MAE as percentages for each ML method. We computed the predictive error for demographic features age, gender, condition plus **(a)** all 53 medicinal features or **(b)** only those medicinal features correlated to BIRB796 at |rs| > 0.3.

### Feature selection and incorporation of genomic information for prediction of drug response

Once we had selected our best “demographic+medicinal” feature set to predict drug inflammatory response, we next investigated if the integration of genomic information into our feature set could provide a predictive advantage, as well as providing further insights into anti-inflammatory drug responses and IBD. After bioinformatic processing and filtering of the patient specific exome capture data based on inter-patient variation, we identified 33,577 SNPs across the 25 patients that were labelled as either homozygous, heterozygous or homozygous ref (see Methods). Adding the SNPs to the demographic and correlated medical features directly resulted in an extremely high dimensional dataset (25 patients × 33,590 features). To counteract the detrimental effect of adding tens of thousands of genomic features describing a relatively small set of patients, we used two approaches for feature selection prior to model training. Firstly, we selected known related gene-centric SNPs using domain insight. More precisely, to select known related SNPs, we used those within genes identified (from target validation.org) as the top ten most associated with Crohn’s and also ulcerative colitis. This resulted in 16 genes (as there was overlap in the top ten between the conditions) and 39 underlying SNPs (see Table S1). Secondly, to incorporate *de novo* insights alongside known SNPs, we used the chi-squared method for feature selection; removing the most independent features compared to the target to predict. In order to define a non-arbitrary cut-off for feature selection with chi2, we sequentially reduced the feature number as input to a model for training and testing, by removing the most independent features in each iteration. We recorded the predictive error and the reduced feature sets for each iteration. Prioritizing reduction of overfitting between the test and training data, the “best” feature set was composed of 40 features (marked with a circular marker on Figure S2). It is also clear from Figure S2 that the sequential removal of SNPs/features improved the predictivity of the initial model by decreasing the predictive error. The 40 reduced features included 32 SNPs (plus 8 medical features). These features were taken forward, combined with the previously defined “known” 39 SNPs and used to train a range of regressors (including hyper tuning) to check if superior accuracy could be achieved with the reduced feature set (see Methods). Note that for the chi2-square feature selection analysis, we used the best model with best hyper-parameters resulting from our previous analysis that incorporated the demographic and correlated medical data. Interestingly, there was no overlap between the 32 SNPs defined by chi2 and the 39 previously “known” SNPs, demonstrating the complementarity of our approaches.

To quantify the benefit of adding genomic features to our existing demographic and medicinal feature set, alongside the impact of feature selection for the SNPs, we again used BIRB796 at 10nM as an example to highlight our comparison of the predictive capability of the derivative models. Figure 4 shows that the addition of curated plus known genomic information gave a lower predictive error generally compared to using the full SNP set. From this analysis, we derived our final best “demographic+medicinal+SNP” feature set that incorporated the demographic features age, gender and condition, correlated medicinal features for BIRB796 and our curated, filtered SNPs. The best model, that gave the lowest predictive error over 10-fold cross validation, was KNN (Figure 4b) with a median MAE of 4.98%. The median and average MAE’s are close to those obtained with KNN (Figure 3b) and our previous best “demographic+medicinal” feature set that did not incorporate genomic information. The incorporation of genomic features produced a slight increase in the average predictive error (0.28%). This was however balanced with an increase in model stability on cross validation, demonstrated by a 0.39% decrease in standard deviation of MAE over 10-fold cross validation. Furthermore, incorporating genomic information clearly had the potential to provide additional insights into the predicted phenotype when we incorporated model explanation in our analysis, as we show in the section below.

**Figure 4.**
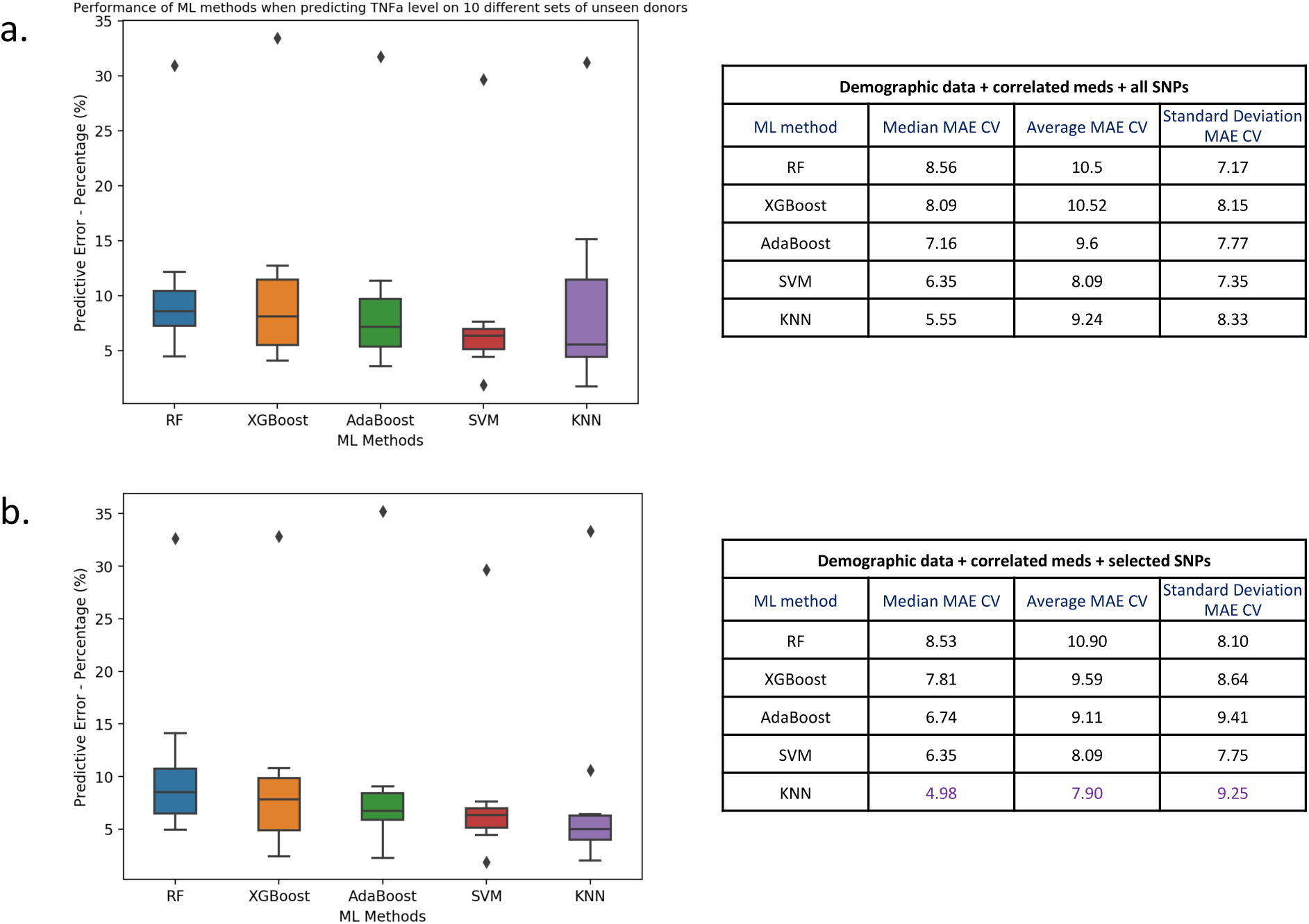
Comparison of ML model error rates for the prediction of BIRB796 (10nM) drug response for different combinations of demographic, medicinal and genomic features. Here we show box plots (left) of mean absolute error values (as percentages) computed during 10-fold cross validation. The horizontal line in each boxplot is the median of the MAE over 10 folds, where each of the test folds has 3 randomly chosen patients. Note that all target drug responses have been normalized on a scale of 0-1 and here we show percentages of MAE values. On the right, we report median, average and standard deviation MAE as percentages for each ML method. We computed the predictive error using demographic features age, gender and condition and correlated medicinal features plus **(a)** all SNPs (33,577) or **(b)** the 71 curated known and associated SNPs.

### Comparing model explanations after the integration of multiple datasets for BIRB796

We generated a series of models as we iteratively added more datasets into our analysis, defining the best model at each stage by considering input features and predictive error on the test dataset, while performing cross validation. We propose however that the predictive error or accuracy is not the only aspect to consider when evaluating a ML model. We highlight the usage of model explanation to validate and interpret the predictions generated by our best model (KNN) trained on our best features sets (“demographic+medicinal” and “demographic+medicinal+SNP”). The explanations of the predictions allowed us to assess the biological insight that can be gained from integrating multiple data sources into the predictions and to instil trust in the results produced by our best models.

To generate model explanations, we applied a state-of-art explainable AI algorithm called SHapley Additive exPlanations (SHAP) [26] to generate the plots shown in Figure 5 (see Methods). Figure 5a and 5b show the ranked lists of “demographic+medical” and “demographic+medical+SNP” features respectively, based on their average absolute SHAP impact value in the predictions generated by KNN for the entire set of 25 donors. For both sets of features, the most impactful feature for the model is condition defined as ulcerative colitis (UC) or Crohn’s. Figure 5c and 5d show, for the “demographic+medical” and “demographic+medical+SNP” features respectively, how each individual feature (each row in the SHAP dot plot) is driving the prediction of a higher or lower TNFα level for each donor (each dot). For the donors (dots) on the right side of the x-axis a positive impact value of a feature drives the prediction of a higher TNFα level, while for the donors on the left side of the x-axis, a negative impact value of a feature drives the prediction of a lower TNFα level. The top 20 features shown here significantly impacted the model’s predictions and the values of some of these features (e.g. condition, gender and a number of SNPs) informatively formed separated clusters (red or blue) that discriminated the positive and negative impact of a feature on the model prediction i.e. respectively high or low TNFα levels.

**Figure 5.**
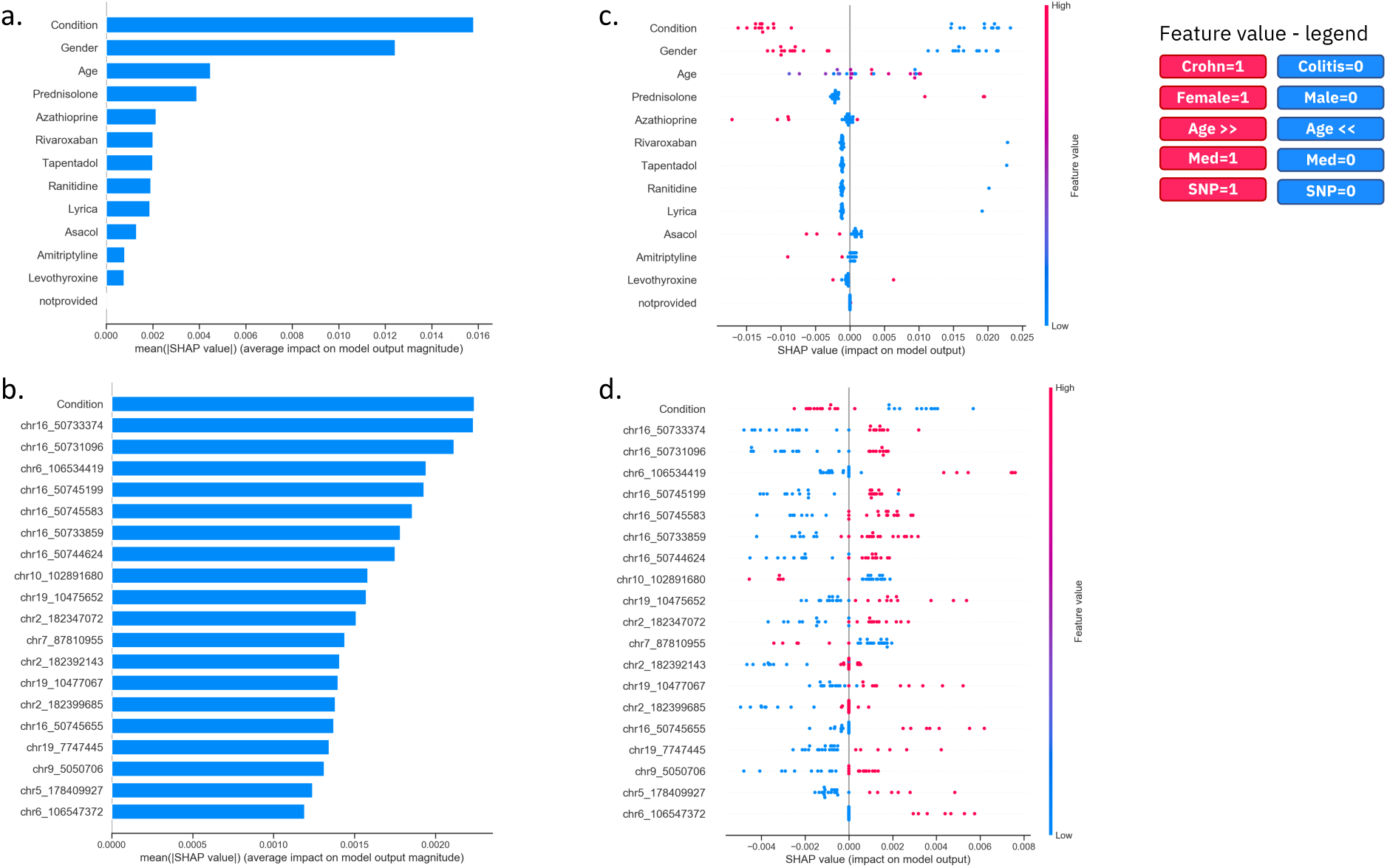
Comparison of ML model explanations for the prediction of BIRB796 (10nM) drug response for demographic, medicinal and genomic features. Here we show SHAP plots that contain explanations for the predictions generated by our best model KNN using **(a, c)** our best “demographic+medicinal” feature set and **(b, d)** our best “demographic+medicinal+SNP” feature set. The SHAP bar plots **(a, b)** show the top 20 features ranked by their impact on the model prediction. The SHAP dot plot **(c, d)** shows the same top 20 ranked features together with the weight of each feature (row) for the prediction of TNFα level for each donor (a donor is a blue or red dot). The figure legend (top right) details the colour and corresponding value that each feature has for each donor (coloured dot). For example, donors that are older are red dots in the plot, while younger donors are blue dots. Similarly, donors that have a SNP allele are shown as red dots, while donors with the reference allele are represented as blue dots.

Focusing firstly on the explanation of our most impactful features from the “demographic+medicinal” model (Figure 5a and 5c), our most impactful feature was “condition (being UC or Crohn’s disease), where we observed a clear separation between patients with Crohn’s and UC. Here, UC patients typically had a higher predicted TNFα level, suggesting that they would be expected to have a lower response to BIRB796 treatment at this concentration. Gender also showed a clear separation of males and females, where females tended towards a higher predicted TNFα level (meaning a lower anti-inflammatory effect of the test drug). We observed a positive correlation between age and SHAP value (impact on model output), indicating that a higher age likely contributes to a prediction of a higher TNFα level or lower response to BIRB796 treatment. We also noticed that some prescribed medicines (Azathioprine, Asacol (5-ASA) and Amitriptyline) routinely contributed to predictions of lower TNFα levels (greater test drug effect), while others (Prednisolone) contributed to the prediction of higher TNFα levels (lower test drug effect). Interestingly, the mechanisms of action of Azathioprine, Asacol, Amitriptyline and Prednisolone have all been linked to MAPK activity as described below.

Azathioprine is a purine prodrug which requires complex conversion to its active metabolites to elicit its intended biological activity as an immunosuppressant via inhibition of purine synthesis. The enzyme GST-M1 is involved in the metabolism of azathioprine [27]. Its expression has been linked to greater azathioprine efficacy [28] but has also been implicated in the adverse effects and toxicity frequently observed in patients who are either on a high dose or on long term treatment with azathioprine [29]. Another role of GST-M1 is as a negative regulator of p38 MAPK by physically sequestering ASK-1, a MAPK kinase kinase, which activates p38 MAPK [30]. Stress triggers, such as pro-inflammatory cytokines, have been shown to promote the dissociation of GST-M1 from ASK-1, thus activating p38 MAPK [30]. Under these inflammatory stresses (which would be analogous to the conditions in our *in vitro* stimulated explant model) it could be expected that BIRB796 (a potent p38 MAPK inhibitor) would show good efficacy in patients who had previously shown a sufficient response to be administered azathioprine chronically.

The mechanism of action of Asacol (5-ASA) is not fully understood; however, some of its biological activity in IBD has been attributed to it being a PPARγ ligand [18]. PPARγ, when activated, acts as a negative regulator of JNK/p38 MAPK signalling [31], both of which BIRB796 is a direct inhibitor of [20]. As 5-ASA (via PPARγ) acts on the same signalling pathway(s) as BIRB796, this could therefore explain why patients who have previously shown efficacy to 5-ASA would also respond favourably to BIRB796.

Amitryptyline is a tri-cyclic antidepressant medication with a mechanism of action which has been linked to inhibition of PKC phosphorylation. Although less characterised in IBD, PKCε has been shown to be an upstream regulator of p38 MAPK in rat models of stress and anxiety [32]. In these model’s amitriptyline has been shown to inhibit phosphorylation of both PKCε and also p38 MAPK. PKCε has also been shown to be a key signalling molecule in the development of bacterial pathogen-induced colitis in human intestinal lineage Caco-2 cell monolayers [33]. As Amitryptyline may share a similar inhibitory action on the p38 MAPK pathway, this may explain why patients who have shown efficacy to Amitryptyline may also show efficacy to BIRB796.

Prednisolone is a glucocorticoid steroid medication commonly prescribed in IBD patients. Glucocorticoid signalling has been closely linked to MAPK signalling pathways, with p38 MAPK shown to play a key role in the expression and sensitivity of glucocorticoid receptors to ligands [34]. Phosphorylation of p38 MAPK has been shown to phosphorylate the glucocorticoid receptor and therefore downregulate its activity [35]. It may therefore be possible that patients who respond well to glucocorticoid therapy such as prednisolone do not have a high level of activated p38 MAPK driving their disease. Such patients would therefore be likely to have a poor response to BIRB796.

Focusing next on the explanation of our most impactful features from the “demographic+medicinal+SNPs” model (Figure 5b and 5d), here, condition was also in the top 20 most impactful features driving the model predictions and it had a directional impact on prediction that was consistent with the “demographic+medicinal” model. Features such as age, gender and medicinal information, on the other hand, featured in the top 20 impactful features for the “demographic+medicinal” model but disappeared here, replaced by more impactful genomic SNP information. In total, 19 of the top 20 most impactful features were genomic SNP features, with many showing a clear separation between patients with different SNP alleles we typically encountered only 2 of 3 possible alleles (e.g., homozygous reference, heterozygous and homozygous alternate) across the 25 patients that formed red and blue clusters accordingly. All SNPs in Figure 5d showed heterozygous SNPs (red) and homozygous reference alleles (blue). There is a strong bias towards cases where homozygous reference alleles drive a lower predicted patient inflammatory response (better response to BIRB796) compared to heterozygous SNPs that contributed to the prediction of a higher response (89.5% of cases). Only two SNPs (ranked ninth and twelfth) showed heterozygous SNPs that are driving the prediction of a lower patient inflammatory response compared to the homozygous reference alleles driving a higher patient inflammatory response.

### Using transcriptomic information as a confidence metric for informative SNPs

Previously we highlighted that 19 of the top 20 most impactful features for our “demographic+medicinal+SNPs” model were genomic SNP features (Figure 5d). Next, we generated RNA-seq information for a subset of the 25 patients to validate the SNPs (see Methods). Firstly, we assessed SNP presence in the RNA-seq data, validating 8 of the 10 exon derived SNPs - with 100% of the 8 SNPs that had sufficient coverage in the RNA-seq data (>5X) being validated (Table 2). The 19 most impactful SNPs were found to affect 9 different genes. As such, we analysed the RNA-seq data for these 9 genes to assess SNP effect on transcript structure and SNP association with gene expression level. From this analysis, all but one (TRAPPC5) of the 9 genes showed an association between the RNA-seq data and at least one of their SNPs, or else a non-synonymous SNP. We observed that 3 of the 9 genes showed correlation |rs| >0.3 between a SNP allele and their gene expression level; ITGA4 with an intron variant, NOD2 with two exon variants and PRDM1 with a 5’UTR variant (Table 2; Figure S3). An additional 2 genes (plus PRDM1 correlated previously) showed correlation |rs| > 0.3 between a SNP allele and the length (in bp) of their longest transcript; ADAM22 with an intron variant, JAK2 with a 5’UTR variant and PRDM1 with one 5’UTR and one missense exon-based variant (Table 2; Figure S3). Many of these SNPs were synonymous SNPs that do not change amino acids but can disrupt transcription [36], splicing [37], co-translational folding [38], mRNA stability [39], and cause a plethora of other functionally relevant changes e.g., altering transcription and splicing regulatory factors within protein coding regions [40], thus potentially modulating gene expression. In addition to missense variants in PRDM1 and NOD2, a further 3 genes of the 9 (TLX1, GRM6 and TYK2) showed non-synonymous or missense changes, which result in changes in the coding for an amino acid and could affect drug metabolism/excretion efficiency or have effects at the level of the target.

**Table 2.**
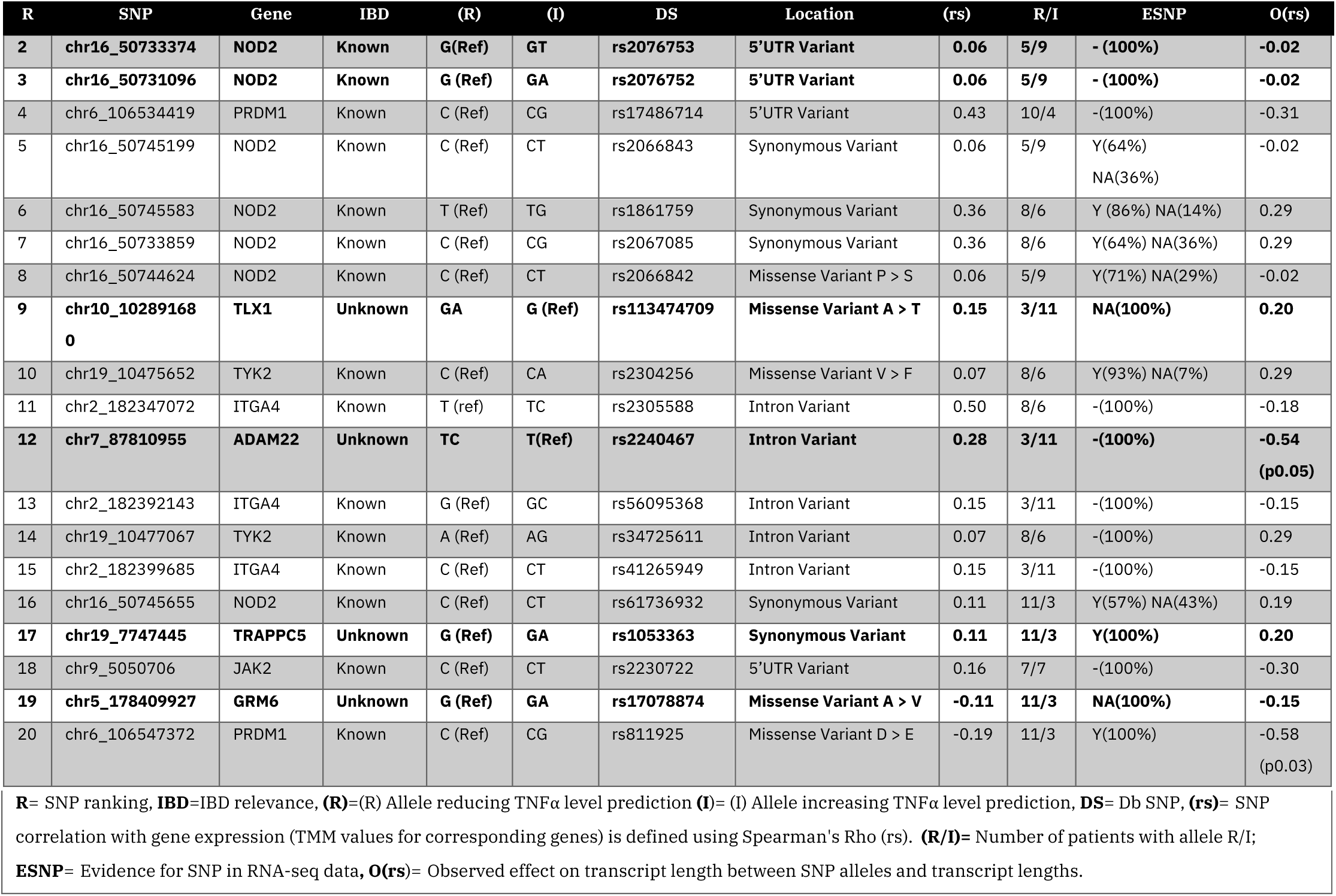
Validation of 19 most impactful SNPs for drug response prediction using RNA-seq data. SNP rank (R) is derived from the model explanation of the best “demographic+medicinal+SNPs” model. Location and demographic association of SNPs is derived from (https://www.ensembl.org/vep) Ensemble Variant effect Predictor. RNA-seq data is available for 14 /25 patients. Evidence for SNP in RNA-seq data (ESNP) is denoted yes (Y) if SNP allele is observed in the appropriate matched samples, no (N) if sufficient coverage is available but no SNP allele is seen, not available (NA) if insufficient sequencing coverage is available (gene not expressed) and “-” is used if the SNP is not in an exon. Observed effect on transcript length (O(rs)) denotes Spearman’s Rho correlation between SNP alleles and transcript lengths of the respective patients from the RNA-seq data. For correlations SNP alleles are converted to 1, while donors with the reference allele are represented 0.

Interestingly, 5 of the 9 genes represented in Table 2 were from our previously “known” list of those highly associated with Crohn’s or UC (Table S1), however the remaining 4 (TLX1, ADAM22, TRAPPC5 and GRM6) were not selected on this basis but their associated SNPs were derived from our chi2 feature selection revealing potential new insights into IBD drug response. Investigation of these genes rationalised their prioritization by the ML model for prediction or discrimination of inflammatory response between patients; TLX1 is linked to colorectal adenocarcinoma that chronic or poorer prognosis IBD patients are known to have a higher risk of developing, ADAM22 has been implicated in lipopolysaccharide-induced inflammation which can stimulate TNFα [41], TRAPPC5 has had family members i.e. TRAPPC9 demonstrating an ability (with NIBP) to potentiate TNFα-induced activation [42] and GRM6 encodes a glutamate receptor which has significance since it has been reported that peripheral glutamate and peripheral glutamate receptors contribute to inflammatory pain [43].

The majority (58%) of the individual 19 SNPs in the top 20 most impactful features for our “demographic+medicinal+SNPs” model, were either non-synonymous variants or synonymous variants that directly correlated with our RNA-seq analysis. However, since multiple SNPs were present for the known Crohn’s and UC genes, we wanted to test if a single representative SNP per gene was sufficient to equal our median model MAE of 4.98% e.g., using the most highly correlated SNP with the RNA-seq or else a non-synonymous SNP. Therefore, we reduced the 71 SNPs down to 43 accordingly (Table S1) and compared the resultant model “demographic+medicinal+filterSNPs”. This refinement did not improve our best median MAE, in fact, it increased to 6.14%, showing that multiple different SNPs, even for a single gene, can be individually informative, if that gene is of significance to the phenotype being predicted, as is the case here.

### Extension of predictive analysis to additional doses and compounds

Initially, as proof of concept we focused on a single compound (BIRB796 and dose 10nM); however, pharmacological data (TNFα response) was available for the three drugs previously mentioned and for prednisolone and BIRB796 across two doses. The doses were chosen based on an approximation of the mean local concentrations expected in patients. As such, we trained additional ML models for each of the remaining compounds and doses (see Table in Figure 6). We curated features for model training as per our investigation of BIRB796 combining the same demographic features used previously, with the medical features that correlated with drug response, per drug (Table 1) and also the 39 known related SNPs from the top ten genes most associated with Crohn’s and UC (Table S1).

**Figure 6.**
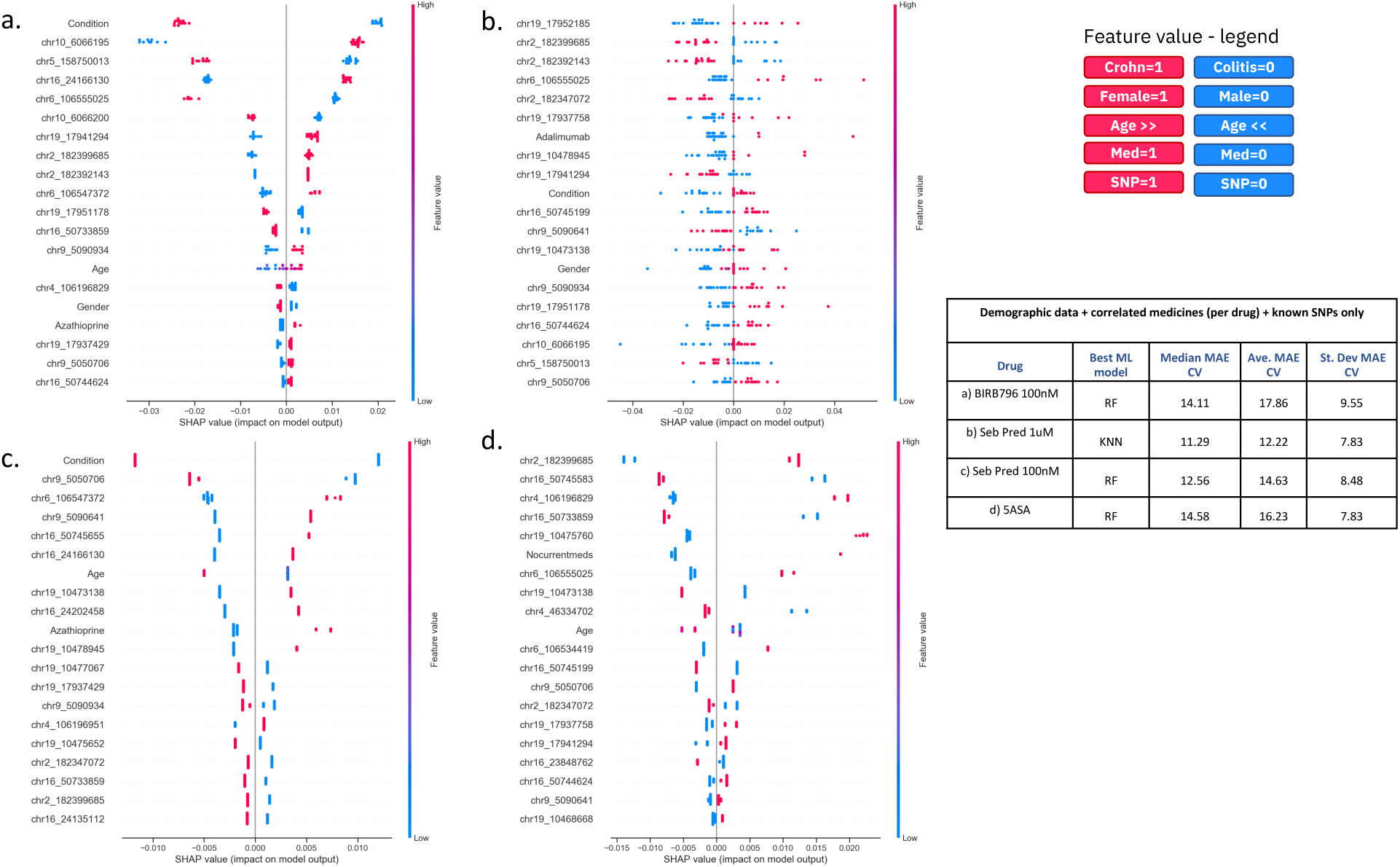
Comparison of ML model explanations for the prediction of response to three drugs (x2 doses) for demographic, medicinal and genomic features. Here we show SHAP plots that contain explanations for the predictions as generated by our best ML models (as defined in the table to the right) using “demographic+medicinal+SNP” features as appropriate per drug for **(a)** BIRB796 100nM **(b)** Pred 1uM **(c)** Pred 100 nM **(d)** 5-ASA. Plots **(a-d)** show the top 20 ranked features together with the weight of each feature (row) for the prediction of TNFα level for each donor (a donor is a blue or red dot). Figure legend details the colour and corresponding value that each feature has for each donor (coloured dot). The table to the right shows, for each drug, the best ML model and the median, average and standard deviation mean absolute error values (MAEs) computed during 10-fold cross validation. Note all target drug responses have been normalized on a scale of 0-1 and here we show percentages of MAE values.

Investigating the explanation of our most impactful features from the best “demographic+medicinal+SNPs” models per drug/dose (Figure 6a-d), we noted that two SNPs were common to all 5 treatments: chr2_182399685 (an intron variant in ITGA4) and chr9_5050706 (a missense variant in JAK2). Additionally, we defined features that were unique to the top 20 most impactful model features for each single compound and dosage (Table S2). BIRB796 at 10nM had the most unique features (6 SNPs highlighted in bold in Table 2) and BIRB at 100nM had the least unique features (1 intron variant rs11256369, in the gene IL2RA). Otherwise, the model for Pred at 1uM had two unique features in its top 20 most impactful, the intron variant (rs3212733) in the gene JAK3 and the prescribed medicine Adalimumab. The model for Pred at 100nM had three unique features in its top 20 most impactful, the synonymous variant (rs3729904) and intron variant (rs1013316) in the gene PRKCB and the missense variant (rs2454206) in TET2. The model for 5-ASA had five unique features in its top 20 most impactful, firstly, the designation that the patient was taking no other medicines, plus four SNPs including two intron variants (rs12720270 and rs12720299) in the gene TYK2, an intron variant (rs279827) in the gene GABRA2 and the intron variant (rs3729883) in the gene PRKCB.

## Conclusions

In this study, we propose a ML workflow to predict inter-patient variation in drug response and we use explainable AI to identify and rank important multi-modal features that guide these predictions. We integrate diverse data sources, including multi-omic, demographic, medicinal (prescribed medicines) and pharmacological data to facilitate precision medicine. We demonstrate the potential of combining preclinical functional characterisation of drug efficacy and inter-patient variation in drug response, with state-of the-art omics, bioinformatics and ML/AI approaches as a new way to model precision medicine strategies at the early stages of drug development.

As an exemplar project, the number of patients was relatively low and findings are therefore tentative; however, this study was also designed to explore the potential for such projects during pre-clinical drug development, where budgets are limited and projects exploring hundreds of patients may be too costly. Nonetheless, our best model was able to predict inflammatory drug response from a combination of integrated demographic, medicinal and genomic features from only 25 patients with an error as low as 4.98% on unseen patients. More importantly, clear variations in drug effectiveness were observed between patients e.g. considering demographic features such as gender, age or condition or previous medication history.

Our preliminary experimental findings (incorporating RNA-seq data) suggest that genetic polymorphisms in our cohort of IBD patients are linked to variation in response to the anti-inflammatory treatment BIRB796 (doramapimod). In particular, firstly, we associated the presence of the alternate allele of the variant rs2240467 in the gene ADAM22 with a significantly shorter transcript length (rs=-0.54, p=0.05), which has the potential to affect downstream functionality. ADAM22 has been implicated in the release of inflammatory cytokines such as TNFa [41] so this transcript truncation supports our observed tendency for the variant allele of rs2240467 to reduce the predicted TNFα level by our ML model. We interpret this as inducing a better response to the test drug. Secondly, the presence of the alternate allele of the variant rs811925 in the gene PRDM1 was also associated with decreased expression of the gene and a significantly shorter transcript length (rs=-0.58, p=0.03). This was coupled with the ML model predicting a higher TNFα level when the alternate allele was present. BLIMP-1 (the protein encoded by PRDM1) is thought to be critical to the maintenance of immune homeostasis [44] and p38 MAPK has been shown to be a positive upstream regulator of BLIMP-1 expression [45]. Since BLIMP-1 lies downstream of p38 MAPK signalling, this may explain why a p38 MAPK inhibitor such as BIRB796 would demonstrate lower efficacy in patients with mutations that already inhibit BLIMP-1 expression or affect its functionality. These two variants (in ADAM22 and PRDM1) were selected for being amongst the top 20 most predictive for our ML model, but also for being strongly associated with the transcriptomic observations (p<0.05), allowing validation. Interestingly, the variant in ADAM22 was derived from our chi2 feature selection while the variant from PRDM1 was from our previously “known” list associated with Crohn’s or UC (Table S1). Similarly, all SNPs that we found most predictive of drug response included a balance between those from our previously “known” list and those derived *de novo* from chi2 feature selection. This highlights the benefit and complementarity of combining domain knowledge with more standard feature selection approaches.

Finally, we were able to apply our workflow to other drugs or dosages. Model explanations revealed some overlap between the most impactful features that each different model was using to make predictions. On the other hand, we found features (mainly composed of genetic SNP marks) that differentiated the models, targeting them to each specific compound of interest.

While a high volume of functional and genomics data was generated, the total number of patients was low for an association study. For this reason, the scientific conclusions made from the study remain tentative. However, we feel that this project serves to demonstrate well the potential to explore patient stratification strategies, at a much earlier stage, by combining fresh tissue pharmacology, demographic metadata, multi-omics and AI. Future work will involve the extension of this analysis to a larger dataset to investigate the wider adoption of our approach and further showcase its impact for precision medicine.

## Methods

### Demographic feature preparation for ML

Demographic features were codified as follows:

1. Age, originally expressed in intervals, was translated into integers in the interval [0,7] as follows
  a. [18-24] -> 0,
  b. [31-36] -> 1,
  c. [37-42] -> 2,
  d. [43-48] -> 3,
  e. [49-54] -> 4,
  f. [60] -> 5,
  g. [66] -> 6,
  h. [72] -> 7
2. Gender, female or male, was translated into 0 and 1 respectively
3. Condition, Ulcerative Colitis or Crohn’s, was translated into 0 and 1 respectively
4. Resection area, colon or ileum, was translated into 0 and 1 respectively
5. Smoking history had inconsistent metadata e.g. non-smoker, unknown, 5 cig per day, previous, less than 20 a day that was manually transformed to either 0 (non-smoker) or 1 (current/previous smoker) so it was removed from the analysis
6. Alcohol history had inconsistent and largely unknown assignments including none, unknown, occasional, <21 units per week, 15 units per week and 8 units per week.

### *Ex vivo* organoculture

Colon or ileum tissue was ethically obtained from 25 patients, clinically diagnosed with IBD in the form of Crohn’s or ulcerative colitis who were undergoing therapeutic resection surgery. Full thickness mucosal biopsies prepared from each tissue were cultured for approximately 18 hours in a humidified incubator (37 °C, 5% CO_2_), in culture media fortified with either test article or test article vehicle. Each test condition was tested in duplicate culture wells in each donor. Levels of TNFα were measured in culture supernatants using a magnetic bead-based assay for the Luminex MAGPIX platform. Each culture supernatant sample was analysed in duplicate and the mean value used in downstream analyses.

### RNA, DNA extraction

DNA was extracted from approximately 10 mg tissue using the PureLink™ Genomic DNA Mini Kit. DNA quality control was performed using the Agilent 2200 TapeStation and the Genomic DNA ScreenTape kit to determine the DNA integrity number (DIN). RNA was extracted from approximately 10 mg of tissue. Tissue was homogenised and total RNA was then extracted using the miRCURY RNA Isolation Kit–Cell & Plant. Absorbance ratios at 260/280 nM and 260/230 nM were determined as indicators of sample yield and purity. Further RNA quality control was performed using the Agilent 2200 TapeStation and the ScreenTape R6K kit to determine the RNA integrity number (RIN).

### Exome and RNA sequencing

#### WES

Targeted next generation sequencing libraries were prepared using the Ion Ampliseq™ Exome RDY Kit and DNA isolated from baseline lung biopsies. Multiplexed PCR was performed to produce barcoded libraries, using 100 ng of input DNA per sample and 10 amplification cycles. The Ion AmpliSeq™ Library Kit Plus and IonXpress™ Barcode Adapters were used in library preparation, according to the manufacturer’s instructions. Final library concentrations were determined by quantitative real time PCR using the Ion Library TaqMan™ Quantitation Kit. Libraries were diluted to 100 pM, and 2 libraries were subsequently pooled in equal amounts for templating on the Ion OneTouch™ 2 System, using the Ion PI™ Hi-Q™ OT2 200 kit. The Ion Proton™ NGS platform was used for sequencing of multiplexed templated libraries, using the Ion PI™ Hi-Q™ Sequencing 200 Kit and the Ion PI™ Chip Kit v3, according to the manufacturer’s instructions.

#### RNAseq

Following QC, mRNA was enriched using oligo(dT) beads. The mRNA was then fragmented randomly in fragmentation buffer, followed by cDNA synthesis using random hexamers and reverse transcriptase. After first-strand synthesis, a custom second-strand synthesis buffer (Illumina) was added with dNTPs, RNase H and Escherichia coli polymerase I to generate the second strand by nick-translation. The final cDNA library was ready after a round of purification, terminal repair, A-tailing, ligation of sequencing adapters, size selection and PCR enrichment. Library concentration was first quantified using a Qubit 2.0 fluorometer (Life Technologies). Insert size was checked on an Agilent 2100 and quantified using quantitative PCR (Q-PCR). Libraries were fed into Illumina machines according to results from library QC and expected data volume.

#### Organoculture bioinformatic analysis

TNFα levels (pg/mL) determined for each patient sample in the organoculture assay, were normalised against the patient sample matched vehicle control group. The individual drug treated biopsy results were calculated as a percentage of the mean vehicle control group results before both drug treated biopsy results were then meaned to provide a single result for each drug treatment per donor sample.

#### Post-processing of the TNFα level

Post-processing of the TNFα data, the TNFα ranges were as follows for 50 µg/mL 5-ASA 50.5-345.0, for 1 µM Prednisolone 36.8-293.8, for 100 nM Prednisolone 11.1-136.1, for 100 nM BIRB796 9.9-94.6 and for 10 nM BIRB796 50.6-369.1. The overall range across the compound set was 9.9-369.1. As such, for each compound the TNFα ranges were normalized to a scale of 0-1 to enable easier comparison between the compounds.

#### Exome sequence bioinformatic analysis

Torrent Mapping Alignment Program was used to provide IonTorrent AmpliSeq exome sequencing data for each patient. Data was provided as a BAM file aligned to genome reference GRCh37. Genotypes called with Torrent Variant Caller were provided as per sample VCF files. Single nucleotide polymorphisms (SNPs) from the VCF files were merged into a multi-sample VCF and VCF files were then filtered to remove low quality SNPs including those with a depth less than 30.

Post-processing of the SNP data, we extracted 102,601 unique SNPs across the 25 patients. For these unique SNPs we extracted the patient-wise information labelling as either; homozygous (using the allele that the patient has if alternate allele freq >0.8), heterozygous (denoted using the reference then the alternate allele if alternate allele freq <=0.8) and finally, homozygous Ref (if no SNP was recorded for that patient at that position). SNPs were filtered for those showing little variation across the 25 patients i.e. those were 20+ of the 25 patients showed the same SNP allele. We also filtered if the reference or alternate allele were not a single A/T/G/C i.e. no indels. This left 33,577 SNPs. For input into ML homozygous, heterozygous and homozygous reference were transformed into a numerical format.

#### RNA-seq bioinformatic analysis

Raw paired-end reads were obtained in FASTQ format (un-stranded) for 14 of the 25 patients. These reads were filtered with Trimmomatic v0.39 [46] for adaptor sequence and also for quality using a 4-base wide sliding window where reads were cut when the average quality per base drops below 15. Reads were dropped if trimmed below 40 bp long. Surviving reads were aligned to the human reference genome (GRCh38) using HISAT2 v2.1.0 [47] with default parameters. Uniquely aligned reads were selected (if mapped in a proper pair) using SAMtools v1.10 [48] and duplicate removal was performed using MarkDuplicates v2.22.0 [49]. Mapping statistics are shown in Table S3. Gene expression levels were quantified using StringTie [50] and the raw expression counts per gene, were subsequently normalised across the 14 patients using EdgeR to generate TMM values [51]. There were 53,779 genes in the dataset, filtering to focus on those represented at a minimum of 1cpm (counts per million) in at least two samples was performed initially to obtain a final analysis set of 19,731 genes.

#### Tuning, training and evaluating machine learning models

We evaluated the application of five state-of-art machine learning models; random forest (RF) [52], XGBoost [53], Support Vector Machines (SVM) [54], K-Nearest Neighbors (KNN) [55] and Adaboost [56]. The first step before tuning and training our models is to standardise our data using scikit-learn sk-learn.preprocessing.StandardScaler() function. We then split our patient set in training set (80% of the entire data) and test set (the remaining 20% of the data). Finally, we normalised the target to predict (TNFα level), as described in section “Post-processing of the TNFα level” of Methods.

The hyperparameters of each ML model are tuned on the training dataset. Hyper-parameter optimization (HPO) consisted of 200 iterations of a random search with 5-fold cross-validation, using the scikit-learn implementation sklearn.model_selection.RandomisedSearchCV(). Each iteration of the random search used a different combination of randomly selected hyper-parameters. Scikit-learn implementation of RF, Adaboost, SVM, KNN (sklearn.ensemble.RandomForestRegressor, sklern.ensemble.AdaBoostRegressor, sklearn.neighbors.KNeightborsRegressor, sklearn.svm.SVR respectively) and the XGBoost implementation from the conda-forge channel <u>((1))</u> were used for this analysis. Table S4 reports the best hyper-parameters selected by HPO for each model and dataset.

Once tuned on the training dataset, the optimised models were trained to make predictions on unseen test data and their predictive performances were compared using the Mean Absolute Error (MAE). We used the scikit-learn implementation of MAE sk-learn.metrics.mean_absolute_error. The MAE is a measure of errors between paired observations of the same phenomena. In this context is a measure of errors between paired true values and predicted values of TNFα level. More precisely, given *X* and *Y* being respectively true and predicted values and *n* the number of patients, MAE is calculated as:

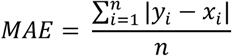

To examine the stability and the generalisability of our models on different randomly selected unseen datasets (i.e., different test sets) we run 10-fold cross validation (10-CV) for each model and each dataset using the scikit-learn implementation sk-learn.model_selection.KFold with parameters n_splits=10 and shuffle=True.

We then computed median, average and standard deviation of MAE across the 10 folds (see Figures 3, 4, and S1 and the table in Figure 6). This allowed us to compare the predictive performances of our models and select the best ML model as the one that provided the lowest MAE on 10-fold cross validation.

#### Explaining the predictions of the best models

Providing explanations for the machine learning model predictions is an important active field of research, as it helps build trust in the models and can provide actionable insights about the target of interest. For this purpose, we used SHapley Additive exPlanations (SHAP) explainability algorithm [26] for its ability to work with any machine learning model. We used the python implementation of SHAP, version 0.35.0, available via the conda-forge channel (https://anaconda.org/conda-forge/shap). SHAP combines game theory with local explanation enabling accurate interpretations on how the model predicted a particular value for a given sample. The explanations are called *local explanations* and reveal subtle changes and interrelations that are otherwise missed when these differences are averaged out. Local explanations allow the inspection of samples that have extreme phenotypes values, e.g. high or low inflammatory response to the drug. By comparing the predictive performances of our ML models, we selected the best model at predicting TNFα level for each feature set and drug. To obtain the appropriate SHAP explainer we combined shap.KernelExplainer with the best hyper-tuned KNN detailed in Table S4. Finally, we used the obtained SHAP explainer to compute SHAP values for the entire set of donors. We then produced the SHAP bar plots in Figure 5a and 5b and the SHAP summary dot plots in Figure 5c and 5d.

## Data Availability

The experimental datasets that were used in this study are available from the ENA Sequence Read archive study PRJEB43220 at https://www.ebi.ac.uk/ena/browser/view/PRJEB43220. Data will be available upon publication.

## Supplemental Figures

**Figure S1.**
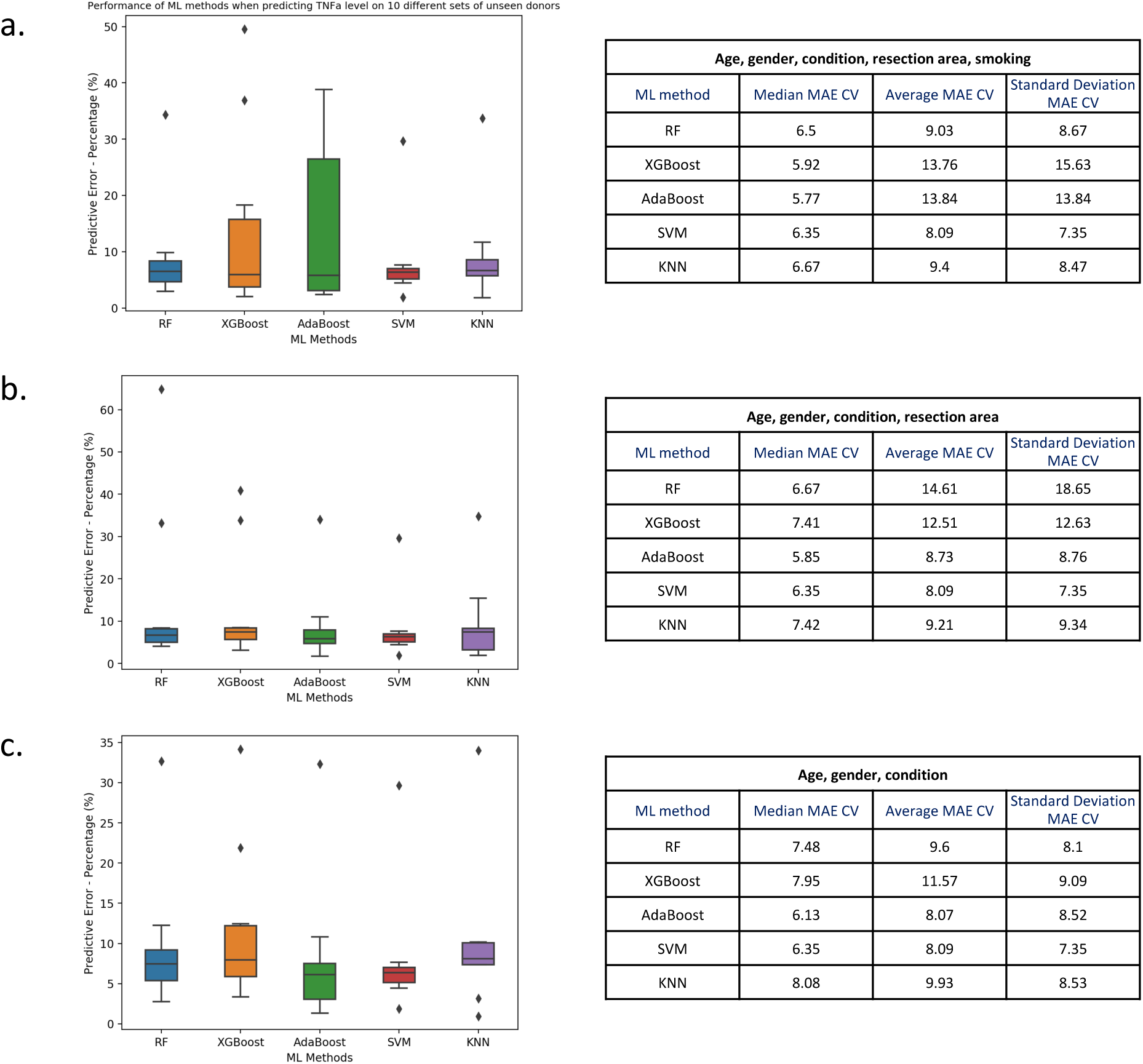
Comparison of ML model error rates for the prediction of BIRB796 (at 10nM) drug response for different combinations of demographic features. Here we show box plots (left) of mean absolute error values (as percentages) computed during 10-fold cross validation. The horizontal line in each boxplot is the median of the MAE over 10 folds, where each of the test folds has ∼3 randomly chosen patients. Note all target drug responses have been normalized on a scale of 0-1 and here we show percentages of MAE values. On the right we report median, average and standard deviation MAE as percentages for each ML method. We computed the predictive error using different combination of demographic features; **(a)** age, gender, condition, resection area and smoking, or **(b)** age, gender, condition, resection area or **(c)** age, gender, condition only.

**Figure S2.**
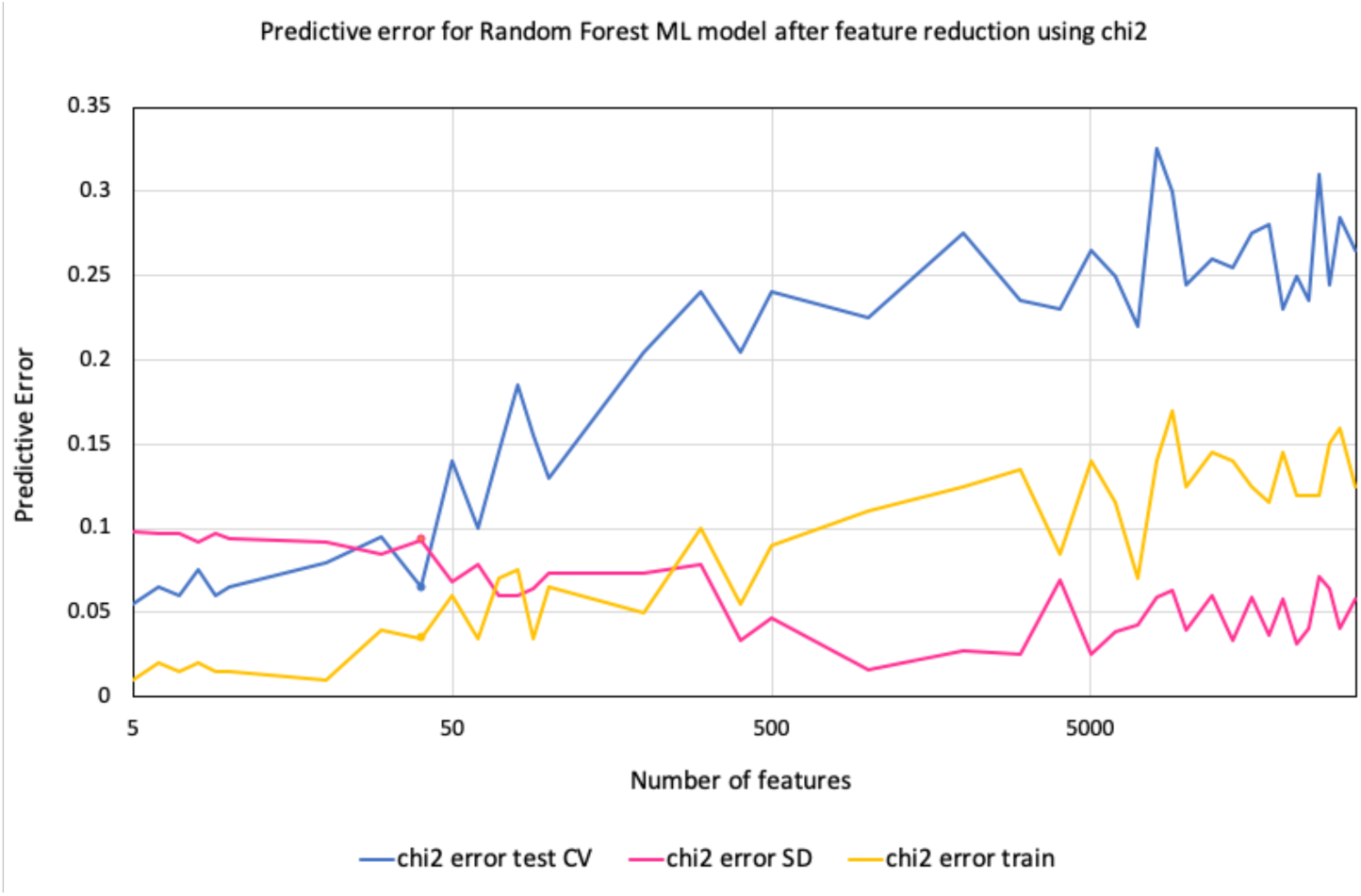
Results of Chi2 test to sequentially remove SNP (genomic) features and observe effect on model MAE rate.

**Figure S3.**
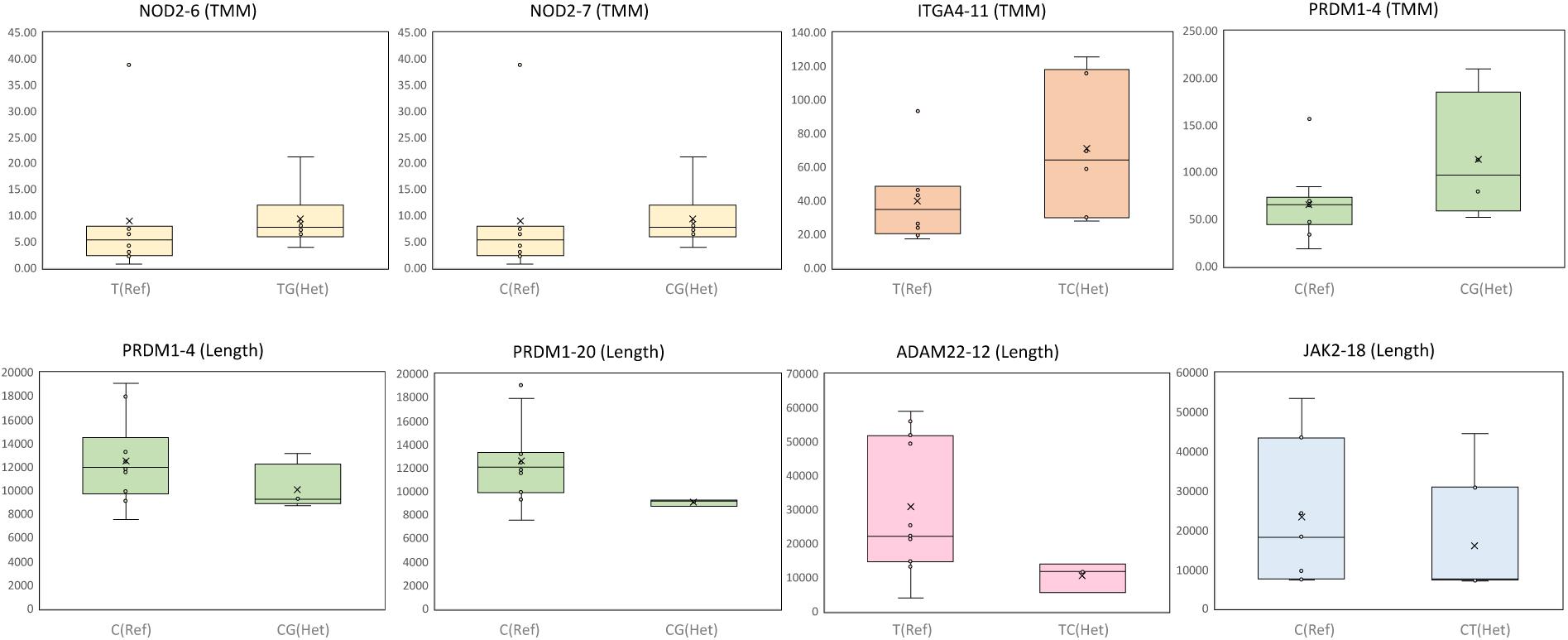
Validation of 19 most impactful genomic SNP features to drug response prediction using RNA-seq data. RNA-seq data available for 14 of the 25 patients. Shown here are the top ranked SNPs from model explanation of best “demographic+medicinal+SNPs” model, if those SNP alleles showed correlation with gene expression (TMM values for corresponding genes) or if they showed correlation with transcript lengths of respective patients using Spearman’s Rho (rs) (> 0.3). Box plots compare the SNP alleles versus their distributions of either TMM values or transcript lengths (bp) for the gene of origin of the SNP. Each box plot is labelled (title) with the “gene of origin of SNP”-“rank of SNP in Table 2” (“if correlated with TMM or Length”). The x-axis denoted the SNP alleles featured in the patient population that are being compared; “Ref” denotes reference allele and “Het” denotes a heterozygous SNP.

## Supplemental Tables

**Table S1.** Top 10 known genes associated with Ulcerative colitis and Crohn’s disease. As identified from target validation.org. Those SNPs selected (one per gene) as non-synonymous or else the most correlated with RNA-seq data are highlighted in bold.

| **Gene** | **Location on GRCh37** | **SNPs within this region** |
| --- | --- | --- |
| **JAK2** | Chromosome 9: 4,984,390-5,129,948 forward strand. | chr9_5090641; chr9_5090934;<br>**chr9_5050706** |
| **NOD2** | Chromosome 16: 50,727,499-50,767,952 forward strand. | chr16_50733859; chr16_50731096;<br>chr16_50733374; **chr16_50744624**;<br>chr16_50745199; chr16_50745583;<br>chr16_50745655 |
| **JAK3** | Chromosome 19: 17,935,589-17,958,880 reverse strand. | **chr19_17937429**; chr19_17937758;<br>chr19_17941294; chr19_17948732;<br>chr19_17951178; chr19_17952185 |
| **TET2** | Chromosome 4: 106,067,032-106,200,973 forward strand. | **chr4_106196829**; chr4_106196951 |
| **GABRA2** | Chromosome 4: 46,245,565-46,477,247 reverse strand. | **chr4_46334702** |
| **IL12B** | Chromosome 5: 158,741,788-158,757,495 reverse strand. | **chr5_158750013** |
| **TYK2** | Chromosome 19: 10,461,205-10,491,248 reverse strand. | chr19_10468668; chr19_10473138;<br>**chr19_10475652**; chr19_10475760;<br>chr19_10477067; chr19_10478945 |
| **PRDM1** | Chromosome 6: 106,441,338-106,557,814 forward strand. | chr6_106534419; **chr6_106547372**;<br>chr6_106555025 |
| **SMAD3** | Chromosome 15: 67,356,101-67,487,507 forward strand. | - |
| **PRKCB** | Chromosome 16: 23,847,304-24,231,932 forward strand. | chr16_24202458; chr16_23848762;<br>chr16_24135112; **chr16_24166130** |
| **IL10** | Chromosome 1: 206,940,947-206,947,886 reverse strand. | - |
| **STAT3** | Chromosome 17: 40,465,342-40,540,500 reverse strand. | - |
| **ITGA4** | Chromosome 2: 182,321,929-182,403,667 forward strand. | **chr2_182347072**; chr2_182392143<br>chr2_182399097; chr2_182399685 |
| **TNF** | Chromosome 6: 31,543,342-31,546,113 forward strand. | - |
| **IL2RA** | Chromosome 10: 6,052,652-6,104,333 reverse strand. | chr10_6066195; **chr10_6066200** |
| **MAPK14** | Chromosome 6: 35,995,454-36,079,013 forward strand | - |

**Table S2.**
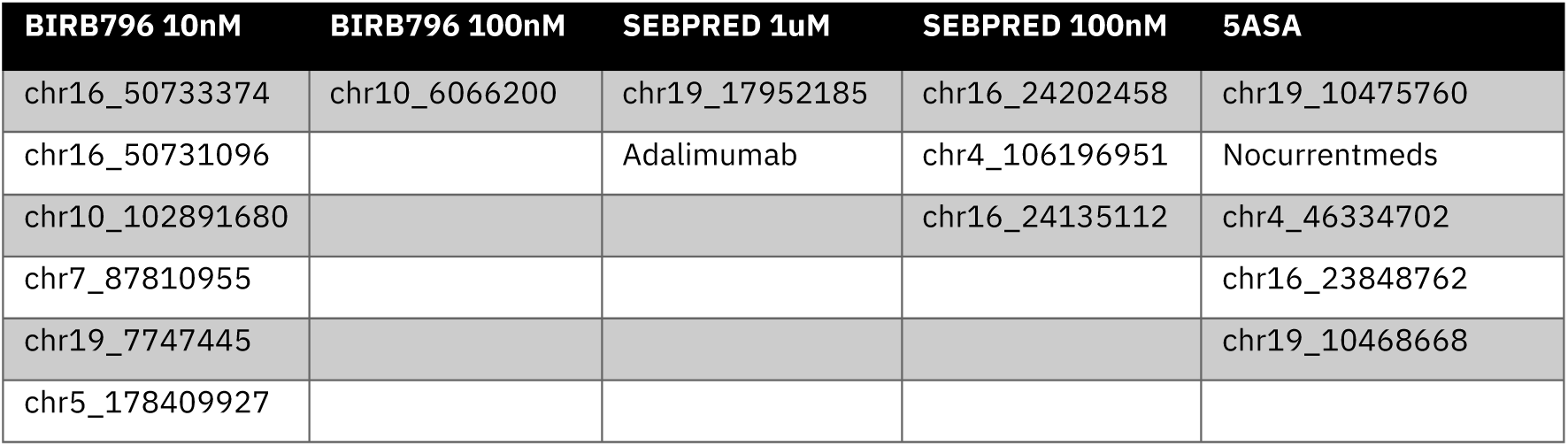
Features specific to the model explanation (among the top 20 most impactful features) of a single tested drug or dosage from the 5 compared in this study.

| BIRB796 10nM | BIRB796 100nM | SEBPRED 1uM | SEBPRED 100nM | 5ASA |
| --- | --- | --- | --- | --- |
| chr16_50733374 | chr10_6066200 | chr19_17952185 | chr16_24202458 | chr19_10475760 |
| chr16_50731096 |  | Adalimumab | chr4_106196951 | Nocurrentmeds |
| chr10_102891680 |  |  | chr16_24135112 | chr4_46334702 |
| chr7_87810955 |  |  |  | chr16_23848762 |
| chr19_7747445 |  |  |  | chr19_10468668 |
| chr5_178409927 |  |  |  |  |

**Table S3.**
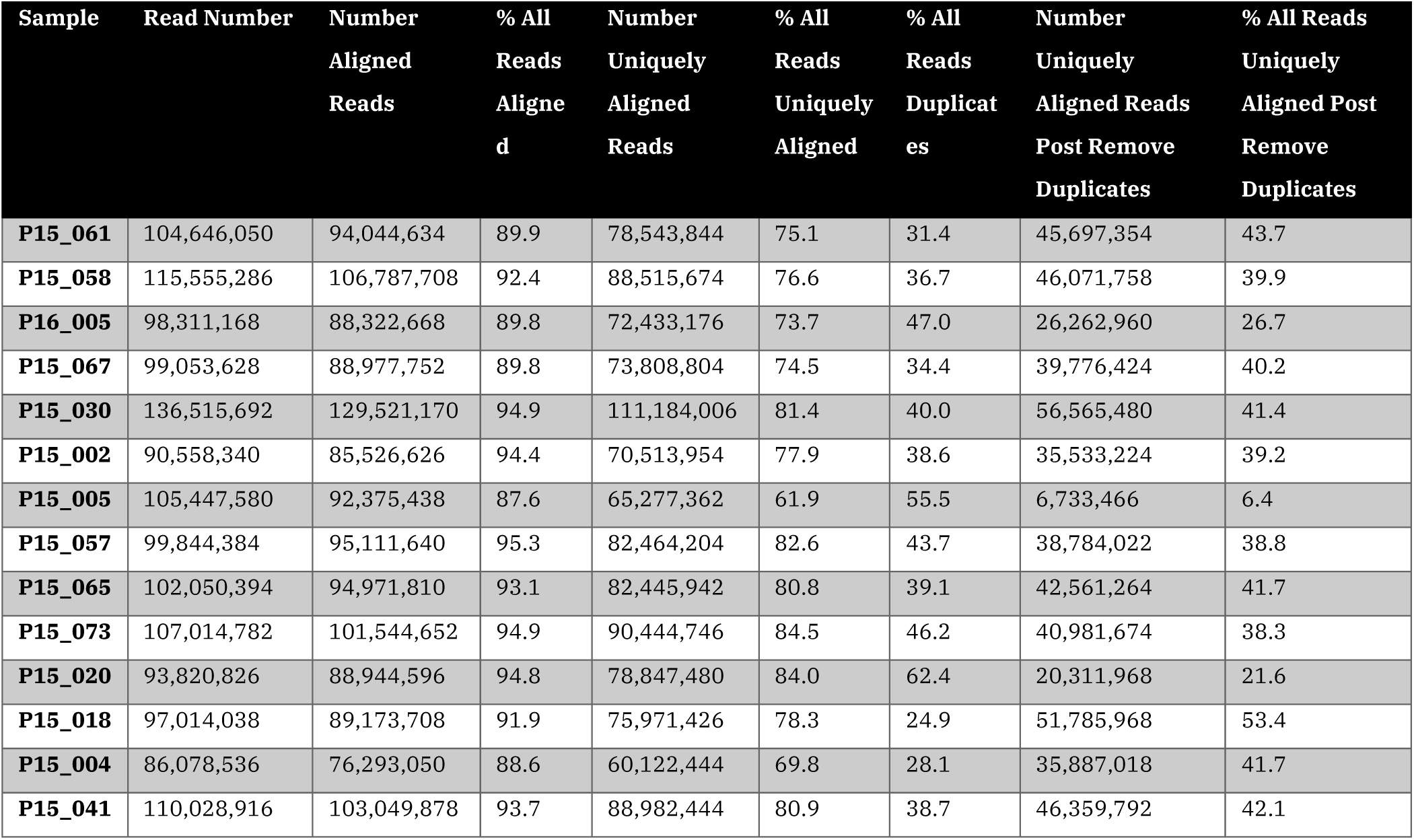
Mapping statistics for the 14 RNA-seq samples. Pre and post alignment.

**Table S4.** Hyper-parameters of the best ML models selected by Hyper-parameter Optimization.

| Drug | Feature set | Best Model | Hyper-parameters |
| --- | --- | --- | --- |
| **BIRB796 10nM** | Demographic + correlated meds | KNN | KNeighborsRegressor(algorithm='auto', leaf_size=30, metric='euclidean', metric_params=None, n_jobs=None, n_neighbors=4, p=2, weights='uniform') |
| **BIRB796 10nM** | Demographic + correlated meds + selected SNPs | KNN | KNeighborsRegressor(algorithm='auto', leaf_size=30, metric='manhattan', metric_params=None, n_jobs=None, n_neighbors=7, p=2, weights='uniform') |
| **BIRB796 100nM** | Demographic + correlated meds (per drug) + known SNPs only | RF | RandomForestRegressor(bootstrap=True, ccp_alpha=0.0, criterion='mse', max_depth=33, max_features='auto', max_leaf_nodes=None, max_samples=None, min_impurity_decrease=0.0, min_impurity_split=None, min_samples_leaf=4, min_samples_split=5, min_weight_fraction_leaf=0.0, n_estimators=72, n_jobs=None, oob_score=False, random_state=42, verbose=0, warm_start=False) |
| **SEBPRED 1uM** | Demographic + correlated meds (per drug) + known SNPs only | KNN | KNeighborsRegressor(algorithm='auto', leaf_size=30, metric='manhattan', metric_params=None, n_jobs=None, n_neighbors=4, p=2, weights='uniform') |
| **SEBPRED 100nM** | Demographic + correlated meds (per drug) + known SNPs only | RF | RandomForestRegressor(bootstrap=True, ccp_alpha=0.0, criterion='mse', max_depth=63, max_features='sqrt', max_leaf_nodes=None, max_samples=None, min_impurity_decrease=0.0, min_impurity_split=None, min_samples_leaf=4, min_samples_split=10, min_weight_fraction_leaf=0.0, n_estimators=40, n_jobs=None, oob_score=False, random_state=42, verbose=0, warm_start=False) |
| **5ASA** | Demographic + correlated meds (per drug) + known SNPs only | RF | RandomForestRegressor(bootstrap=True, ccp_alpha=0.0, criterion='mse', max_depth=40, max_features='sqrt', max_leaf_nodes=None, max_samples=None, min_impurity_decrease=0.0, min_impurity_split=None, min_samples_leaf=2, min_samples_split=10, min_weight_fraction_leaf=0.0, n_estimators=26, n_jobs=None, oob_score=False, random_state=42, verbose=0, warm_start=False) |

## Declarations

### Ethics approval and consent to participate

The West of Scotland Research Ethics Committee (12/ws/0069) granted approval for this study, and patients provided written consent complying with the declaration of Helsinki.

### Consent for publication

All authors have consented to publication of the manuscript. No additional consent required.

### Availability of data and materials

The experimental datasets that were used in this study are available from the ENA Sequence Read archive study PRJEB43220 at https://www.ebi.ac.uk/ena/browser/view/PRJEB43220.

### Authors’ contributions

LJG – Conceptualization, Data Analysis, Writing – all stages; APC - Conceptualization, Data Analysis, Writing – all stages; GM – Conceptualization, Supervision, Writing – original draft, Writing – review & editing; DCB – Conceptualization, Supervision, Writing – review & editing; KB – Conceptualization, Supervision, Writing – review & editing; MM – Supervision; EPK - Supervision, Writing – review & editing

## Acknowledgements

Not Applicable

